# Metabolic expenditure, neurodevelopment, and weight gain into early childhood after fetal growth restriction

**DOI:** 10.1101/2025.03.03.25323221

**Authors:** Cigdem Gelegen, Bella Copley, Neelum Mistry, Chiara Sacchi, Chiara Nosarti, Lorenzo Fabrizi, Anna L David, Kimberley Whitehead

**Author notes:** Contributed equally (Cigdem Gelegen, Bella Copley, Neelum Mistry).

## Abstract

Fetal growth restriction (FGR) subjects exhibit altered metabolism, with higher metabolic rate due to their small body mass, and by adopting strategies to minimise energy expenditure. We investigated how these metabolic differences develop, or manifest in growth trajectories, after FGR, small for gestational age (SGA) (constitutionally small), and normal pregnancies.

We curated a unique composite dataset of 1934 subjects between 14 weeks of gestation and 5 years of age. First, we assessed fetal and infant heart rate to assess whether higher metabolic rate persisted postnatally after FGR. Next, as the largest energy expenditure is brain synaptic maintenance, we tested whether FGR infants had lower white matter volume (proxy for synapse number). Finally, we modelled longitudinal body weight into childhood in FGR, SGA, and control groups, and tested for associations with neurodevelopmental scores at 1-2 years.

Heart rate at rest was higher in FGR fetuses and infants (688 subjects), and FGR infants exhibited a blunted capacity to increase heart rate to a nociceptive procedure (i.e. a physiological challenge). FGR infants had smaller white matter volume (270 subjects). Finally, the more an individual’s weight gain deviated below average curves (1714 subjects), the lower were their motor and cognitive scores at 1-2 years.

## 1. Introduction

Early growth is a metabolically expensive process ^1,2^, with a dramatic peak in metabolic rate to 6 months of age ^3^. The energy-demanding brain is a major driver, particularly as it occupies a larger proportion of total body mass in young children ^4,5^. Whole-body metabolic rate can be inferred from surrogate measures which index resource supply, including heart rate ^6,7^, allowing data to be modelled through gestation when direct measurements of metabolism are impossible.

Antenatally, when metabolic demands are not met by placentally-mediated resources, the result is a fetus small for gestational age (SGA) with evidence of fetal growth restriction (FGR). FGR is a risk factor for stillbirth, and adverse neurodevelopmental and metabolic outcomes in survivors both in the short-term and long-term into adulthood ^8^. Whole-body metabolic rate is higher in FGR ^9–11^ owing to their lower body mass ^12^, with higher heart rates observed in FGR fetuses ^13–16^ and FGR neonates on the first postnatal day ^17,18^. However, it is unknown how heart rate differences develop postnatally, during the crucial first extrauterine weeks after a growth-restricted pregnancy. This is especially important because higher basal heart rate means that this organ is already functioning close to its maximal capacity, which may impair autonomic heart rate reactivity to physiological challenges ^6,19^.

Being solely dependent on fetoplacental nutrient and oxygen exchange to fulfil metabolic demands, FGR fetuses adopt mechanisms to minimise energy expenditure and facilitate growth, including reduced movements and reduced rapid eye movement sleep ^20–22^. A huge energy expenditure during early development is the maintenance of synapses, including the spontaneous electrical activity which sustains them ^23,24^. Post-mortem data demonstrate a decreased number of synapses after FGR ^25–27^. Given that a proxy for synapse number is white matter volume ^28^, we tested here whether neonates with FGR had lower white matter volume (using MRI) or post-synaptic activity (using EEG), which would indicate efforts to reduce brain-related energy costs.

Finally, the degree to which energy costs are met by metabolic rates can be assessed longitudinally using body weight growth trajectories: poor growth indicates a shortfall. Slow fetal growth and lower birth weight than expected for placenta weight indicate placental insufficiency ^29^ and predict later adverse neurodevelopment ^30^. However, such early measurement windows miss the crucial metabolically-demanding postnatal growth phase to 6 months ^3^, when physiological resources, such as iron, are especially easily depleted ^31^. This is when growth falters the most in another vulnerable group (infants living in countries with high rates of undernutrition) ^32^. Here we tracked longitudinal fetal-postnatal growth from 14 weeks since conception to 5 years of age, to assess whether shortfalls in growth during this sensitive period were associated with adverse neurodevelopmental outcomes.

## 2. Methods

### Data

A composite dataset was created. We merged data collected at University College London Hospitals (UCLH) within a neurophysiological research programme ^33,34^, with the Europe-wide (including UCLH) EVERREST cohort ^35^, the Evaluation of Preterm Imaging Study (ePrime; EudraCT 2009-011602-42) ^36^, and open-access data from FEMINA2 ^21^, the IEEE dataport ^37^, and the Norway-Alabama Fetal Growth Study ^38–41^. Of the three open-access datasets, the first two are freely available, while the Norway-Alabama dataset is obtainable upon written application to the Biospecimen Repository Access and Data Sharing (BRADS) committee, Division of Intramural Population Health Research (DIPHR), and permissions were granted to KW in 2023 (https://dash.nichd.nih.gov/resource/LinksToOtherArchives). For all studies, ethical approval to conduct the work was secured from the relevant body (e.g. the NHS Health Research Authority in England) and informed written parental consent was provided.

### Subjects

Being small for gestational age (SGA), defined as estimated fetal weight (EFW) or birth weight <10^th^ centile ^42^, is a risk factor for adverse outcomes, but many SGA subjects are *constitutionally* small and therefore healthy, so risk increases in the case of FGR ^30,43–46^. In FGR, the fetus is SGA with evidence of placental insufficiency ^47^. FGR can be subcategorised into early-onset (FGR-EO) when it is diagnosed before 32 weeks of gestation, which has worse outcomes compared to late-onset FGR which presents from 32 weeks. Consequently, relative to i) controls, being ii) SGA, iii) FGR, or iv) FGR-EO comprises a scale of ascending risk.

Here, controls were defined as live-born infants ≥15^th^ birth weight centile. SGA was defined as <10^th^ birth weight centile, in the absence of meeting criteria for FGR. Where possible, FGR was defined according to the international Delphi consensus criteria ^48^. In the ePrime and Norway-Alabama studies, FGR was defined as <10^th^ fetal or birth weight centile alongside evidence of antenatal adversity associated with placental insufficiency (e.g. maternal preeclampsia). In FGR-EO, before 32 weeks of gestation the fetus is either very small (<3^rd^ centile) or presents with evidence of placental insufficiency including abnormal uterine or umbilical artery doppler measurements ^48–52^. Here, FGR-EO could be confirmed according to these Delphi criteria in 87% of cases (see Table 1), and this category was otherwise assigned to infants with FGR born very preterm (given that they must by definition have FGR-EO). As it was not possible to establish whether FGR was early-onset in some datasets, subjects were assigned to one of two subcategories: FGR (no positive evidence for early-onset) and FGR-EO (positive evidence for early-onset) (Table 1; Table S1).

**Table 1:** Demographics.

|  | Total | Controls | SGA | FGR | FGR-EO<sup>a</sup> |
| --- | --- | --- | --- | --- | --- |
| **No. subjects** | 1934 | 1169 | 309 | 214 | 242 |
| **Sex (% F)** |  | 47.2 | 51.0 | 54.7 | 53.1 |
| **Multiple gestation (%)** |  | 10.8 | 2.6 | 5.1 | 7.0 |
| **Median weight at delivery (g) (interquartile range)** |  | 3120 (2039-3545) | 2570 (2336-2749) | 2350 (2045-2620) | 1052 (748-1515) |
| **Live born (%)** |  | 100 | 100 | 100 | 83.6 |

<sup>a</sup> We were able to confirm in 210/242 (87%) FGR-EO subjects that, before 32 weeks since conception, the estimated fetal weight (EFW) or abdominal circumference (AC) was <3<sup>rd</sup> centile (183 subjects) and/or the umbilical artery had absent end-diastolic flow or the EFW/AC was <10<sup>th</sup> centile with abnormal uterine or umbilical artery doppler measurements (88 subjects) <sup>48</sup>.

Multiple pregnancies were eligible, including in the FGR group so long as only one fetus was small (implying selective FGR, with evidence of discrepancy of umbilical artery doppler measurements between fetuses when available). Exclusion criteria included confirmed or suspected congenital or other non-placentally mediated cause for small fetal size (e.g. structural anomaly, aneuploidy, cytomegalovirus infection). In subjects who suffered an acute neurological insult such as hypoxic-ischemic encephalopathy, data prior to but not subsequent to the insult were eligible for analysis.

### Heart rate

Fetal heart rate at rest was sourced from that calculated during antenatal cardiotocography (CTG) monitoring. Data during labour were not included. Postnatal heart rate was calculated from transcutaneous pulse oximetry or electrocardiography (ECG) recordings. For the latter, automated beat detection was performed using LabChart v.8 HRV software (ADInstruments, Spechbach, Germany), following which all data were visually inspected and missing beats manually added if necessary.

In the UCLH cohort, studies had often been timed around a clinically necessary nociceptive procedure (heel lance) ^34^, and data were separated into pre- and post-procedure. Only pre-procedure data were used to calculate heart rate at rest. To examine heart rate *reactivity*, to this nociceptive procedure, we subtracted the mean heart rate during the preceding 15 seconds, from the maximum heart rate across the subsequent 30 seconds, using pulse oximetry recordings (sampling rate: 2 s).

### Neonatal structural MRI

MRI data analysed were from the ePrime cohort described in ^45^, but our sample size was slightly smaller because we aligned inclusion/exclusion criteria to our own (e.g. that control infants must have a birth weight ≥15^th^ centile (the criteria in ^45^ was ≥10^th^)). Acquisition details are available in ^45^, and the pre-processing methodology used is described in ^53^. In brief, the motion-corrected, reconstructed T2-weighted image was first bias-corrected and brain-extracted. Following this, the brain image was segmented into different tissue types using the validated Draw-EM algorithm ^53^. Segments included white matter, which is analysed here. White matter volume was normalised by total intracranial volume (which included total brain volume and cerebrospinal fluid).

### Neonatal EEG

EEG acquisition details are available in ^34^. EEG during rapid eye movement sleep was analysed, because this is the most prevalent state in neonates ^33^. Data were referred to the midline central channel (Cz), and artefactual segments manually rejected by visual inspection, using EEGLAB v.13 ^54^. The right and left mid-temporal channels (T8 and T7) were available in all recordings, and are the foci of a developmentally-specific spontaneous electrical activity pattern termed temporal theta bursts ^55^. These activity bursts were detected using an algorithm adapted from ^55^. EEG theta activity at T8 and T7 was extracted with a third order Butterworth band-pass filter (4-7.5 Hz). The envelope of the resulting signal was calculated using the Hilbert Transform, and temporal theta bursts were identified as periods lasting 0.3–1.6 seconds, during which the amplitude was more than five times larger than the average amplitude of the signal ^56^. Burst rate and magnitude from T8 and T7 were averaged, after confirming that there were no significant differences.

### Biometry

Estimated fetal weight was calculated using fetal ultrasound biometric measurements according to established formulae ^57,58^, in pregnancies ≥14 weeks since conception ^59–61^. Postnatally, weight was measured directly. We correlated birth weight with placenta weight (trimmed of the umbilical cord and fetal membranes) in a subset of pregnancies ^41^.

### Bayley Scales of Infant and Toddler Development

Bayley scales are age-standardised, and the scores have a reported mean and standard deviation of 100 and 15 respectively. We analysed scores at either 1 or 2 years, utilising the later age point when scores at both ages were available. Cognitive and motor scores were assessed, because the language score was less consistently available. The Bayley scales are periodically updated and the version used differed within this composite dataset (cognitive: Mental Developmental Index or Cognitive Composite; motor: Psychomotor Developmental Index or Motor Composite). This was adjusted for by using a conversion formula so that all scores were aligned to the Cognitive ^62^ or Motor Composite^63^.

### Statistical analysis

Data were processed using SPSS v.29 and R v.4.4 statistical software ^64^. To visualise data, we conducted local polynomial regression fitting of the means. To analyse data, single measures were analysed using a standard linear model, while repeated measures were handled with a multilevel linear model using R package lme4 ^65^. In a multilevel model, the variance of the dependent variable is partitioned into its between-subjects portion (fixed effects: regression coefficients) and its within-subjects portion (random effects: individual deviations in intercept from fixed effects). This ensures that standard errors are valid, as the non-independence of repeated measures is accounted for. Each subject has just one random effect, irrespective of how many measures they had, as this reflects their individual regression line and considers all observed points. When data followed a notably non-linear trajectory, we applied piecewise regression, because local fitting (used for visualisation purposes) does not produce a readily interpretable change curve equation. Piecewise linear regression models allow to fit pieces of a trajectory separately, approximating a curve ^66^, and have been used successfully to model developmental trajectories ^67^. We ‘anchored’ model intercepts at relevant developmental points (e.g. in a model of data from 14 weeks since conception onwards, we might anchor the intercept at the median age at measure in order to report differences between groups at that point, rather than the default first time point of 14 weeks) ^66^. Throughout, we follow an effect size approach to statistical inference, in which t values (the ratio of an independent variable’s coefficient divided by its standard error) larger than 1.96 are reported ^66^.

## 3. Results

The demographics of the subjects are described in Table 1.

### 3.1 FGR-EO subjects had a higher heart rate at rest, and exhibited a blunted capacity to increase heart rate to a physiological challenge

We analysed heart rate at rest between 16 and 92 weeks since conception (approximately 2 months old) (688 subjects, 1218 measures). FGR-EO (but not FGR or SGA) subjects had a higher heart rate than controls (t = 1.982). For example, with the intercept anchored at 32 weeks since conception (median age at measure), their mean heart rate was 3.5 beats/minute higher. Heart rate declined by −0.6 beats per minute/week (Fig. 1a; t = −4.262), which did not interact with group, i.e. the higher heart rate in the FGR-EO group persisted through the developmental period examined with no evidence of this difference closing, after adjusting for sex, antenatal vs postnatal at measure, and data source (FGR-EO: 138 subjects, 644 measures; FGR: 128 subjects, 143 measures; SGA: 26 subjects (no repeated measures); controls: 396 subjects, 405 measures).

**Fig. 1:**
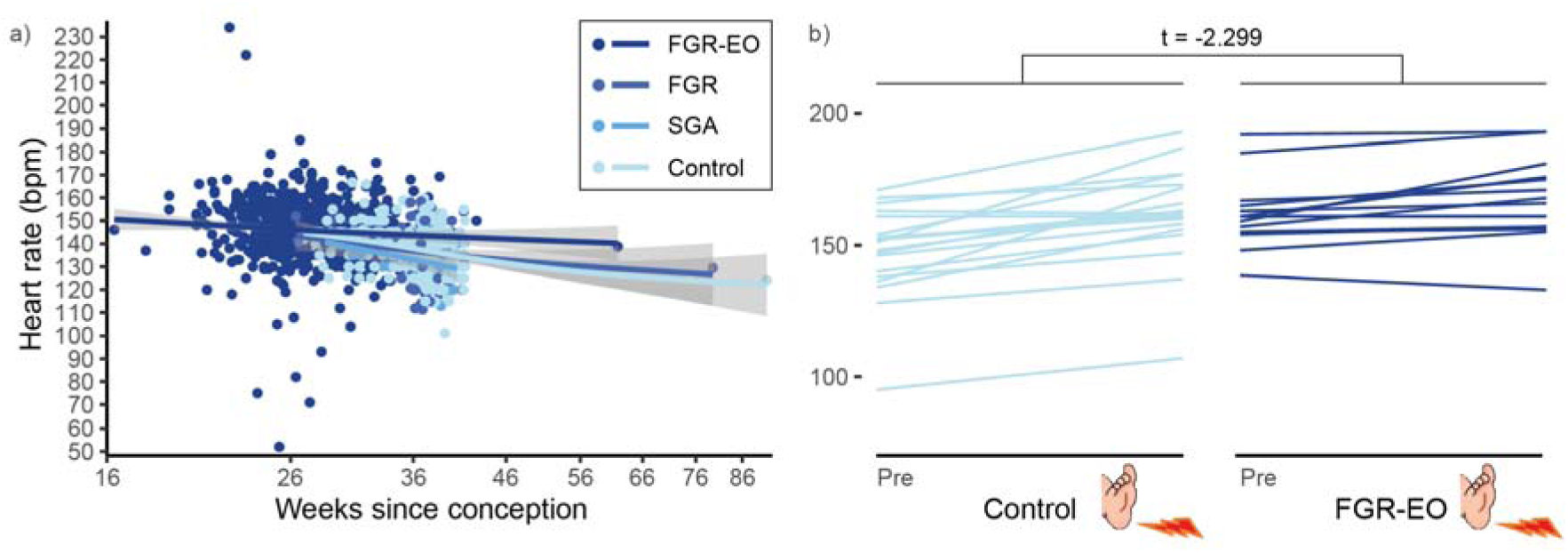
FGR-EO subjects had a higher heart rate at rest than controls, which did not interact with its slope of developmental decline (a), and exhibited a blunted capacity to increase heart rate to a physiological challenge (clinically necessary heel lance) (b). In a) grey shading indicates standard error of the mean; note that the x axis is logarithmic.

Next, we assessed whether heart rate *reactivity* differed in FGR-EO neonates, to a physiological challenge (clinically necessary nociceptive procedure (heel lance)). FGR-EO neonates exhibited a smaller heart rate increase to the challenge after adjusting for sex (Fig. 1b; mean 8 beats lower than controls, t = −2.299, 19 control and 14 FGR-EO subjects matched by weeks+days since conception at birth (median 31+6 vs 31+1) and measure (median 34+2, range 30+6-38+5 vs 33+4, range 30+4-39+3)).

### 3.2 FGR-EO infants had smaller relative white matter volume

Structural MRI data were analysed from very preterm infants (median 29 weeks since conception at delivery (interquartile range 27-31)) scanned between 38-45 weeks since conception. FGR-EO subjects had lower white matter volume than controls (t = −4.065), after adjusting for sex, weeks since conception at delivery, weeks since conception at MRI, and total intracranial volume (FGR-EO: 47 subjects; controls: 223 subjects). In sum, FGR-EO subjects had less white matter than would be expected, even for their small head size (Fig. 2).

**Fig. 2:**
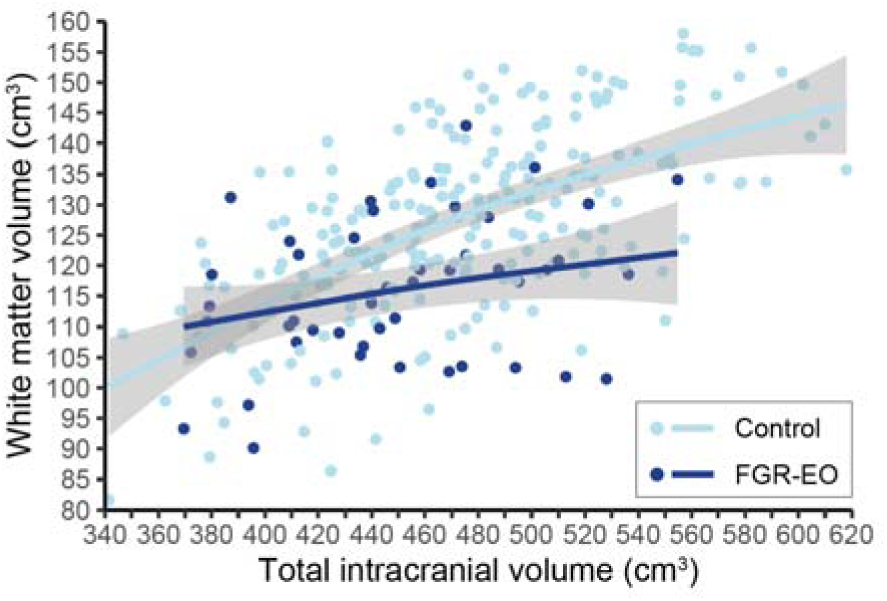
FGR-EO infants had smaller white matter volume. For illustrative purposes, white matter volume is plotted against total intracranial volume, to show the divergence in their relationship between FGR-EO and control infants. Grey shading indicates standard error of the mean.

### 3.3 Temporal theta EEG activity was not different in FGR infants

In line with the literature, temporal theta activity bursts were apparent, coupled to underlying slow activity (Fig. 3a) ^55^. Two developmental phases of its incidence were apparent: a rapid decline to approximately 36 weeks since conception, and a plateau from this point (Fig. 3b). We therefore built a piecewise linear model accounting for this switch point. The t value for the switch point at 36 weeks since conception was large, indicating the need for the piecewise regression (t = 4.956).

**Fig. 3.**
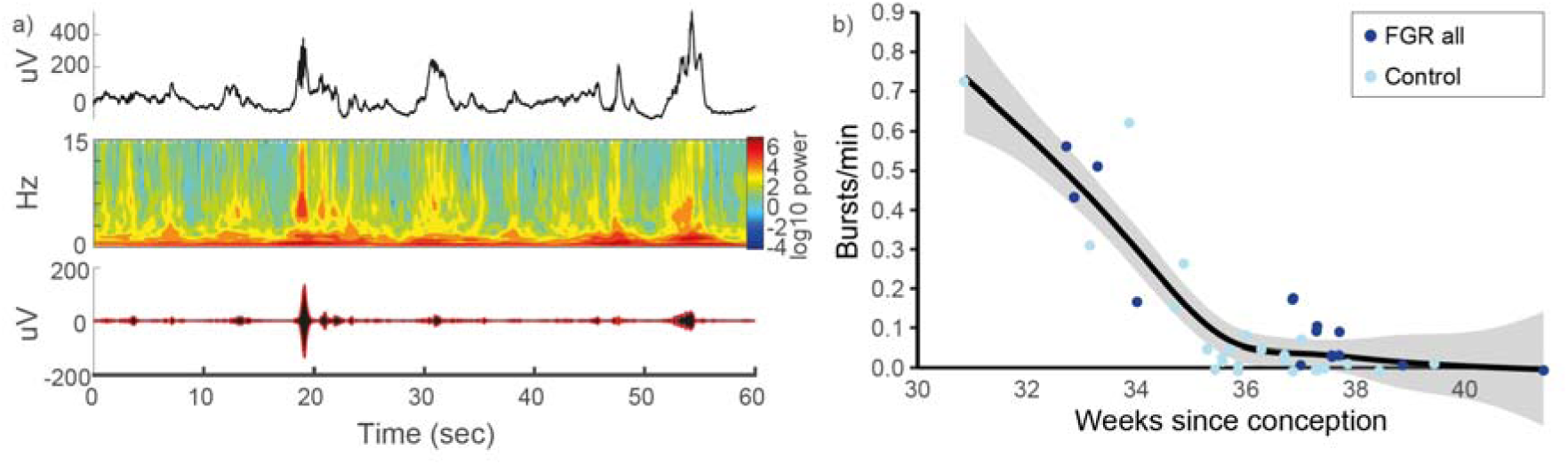
Temporal theta EEG activity was not different in FGR infants. **a)** Upper and middle panels show example temporal theta activity, coupled to underlying slow activity, in the time- and time-frequency domains. Bottom panel shows the Hilbert Transform applied to extract bursts. b) Temporal theta burst rate in controls and FGR (owing to the relatively small sample size, FGR-EO and FGR subjects are pooled into ‘FGR all’ (the colour allocated is that used for FGR-EO in other plots, given that the ratio of FGR-EO: FGR was 11:5)). Data points are plotted slightly jittered to visualise overlapping values. As there was no difference between groups, data were pooled for the local polynomial regression fitting. Grey shading indicates standard error of the mean.

Neither temporal theta burst rate (Fig. 3b) or its magnitude (data not shown) differed in the FGR group, nor was there an interaction between its developmental decline and group, after adjusting for sex and weeks since conception at delivery (25 control and 16 FGR subjects).

### 3.4 FGR subjects had lower birth weight than expected for placenta weight

After showing that FGR subjects had to work harder to meet metabolic demands, and exhibited strategies to reduce energy costs, we next assessed body weight: lower body weight than expected indicates a shortfall between energy costs and metabolic resource supply.

We began by modelling weight at live birth, which is a proxy for fetal growth, and assessed its relationship with placenta weight in a group of 391 subjects delivered between 27-43 weeks since conception (343 controls, 36 FGR, 10 FGR-EO, 2 SGA). Birth weight increased with placenta weight overall (t = 10.421), but was 638 and 784 grams lighter than expected for placenta weight in the FGR and FGR-EO groups relative to controls (t = −8.596 and t = −4.037 (too few SGA subjects to compare)), after adjusting for sex, weeks since conception at delivery, and data source.

### 3.5 FGR and SGA subjects continued to have lower body weight to five years of age

We went on to model body weight longitudinally, from the antenatal period (estimated fetal weight) through the postnatal period (actual weight), across groups. Between 14 weeks since conception and 5 years, three developmental phases of weight gain were apparent: relatively modest weight gain to approximately 25 weeks since conception, a subsequent acceleration to 70 weeks since conception, and then a deceleration from this point (approximately 6 months old) (Fig. 4). The t values for the switch points at 25 and 70 weeks since conception were large, indicating the need for the piecewise regression, and their positive and negative signs confirmed acceleration and then deceleration of weight gain respectively (Fig. 4; t = 21.14, t = −161.97; 1714 subjects, 7454 measures).

**Fig. 4.**
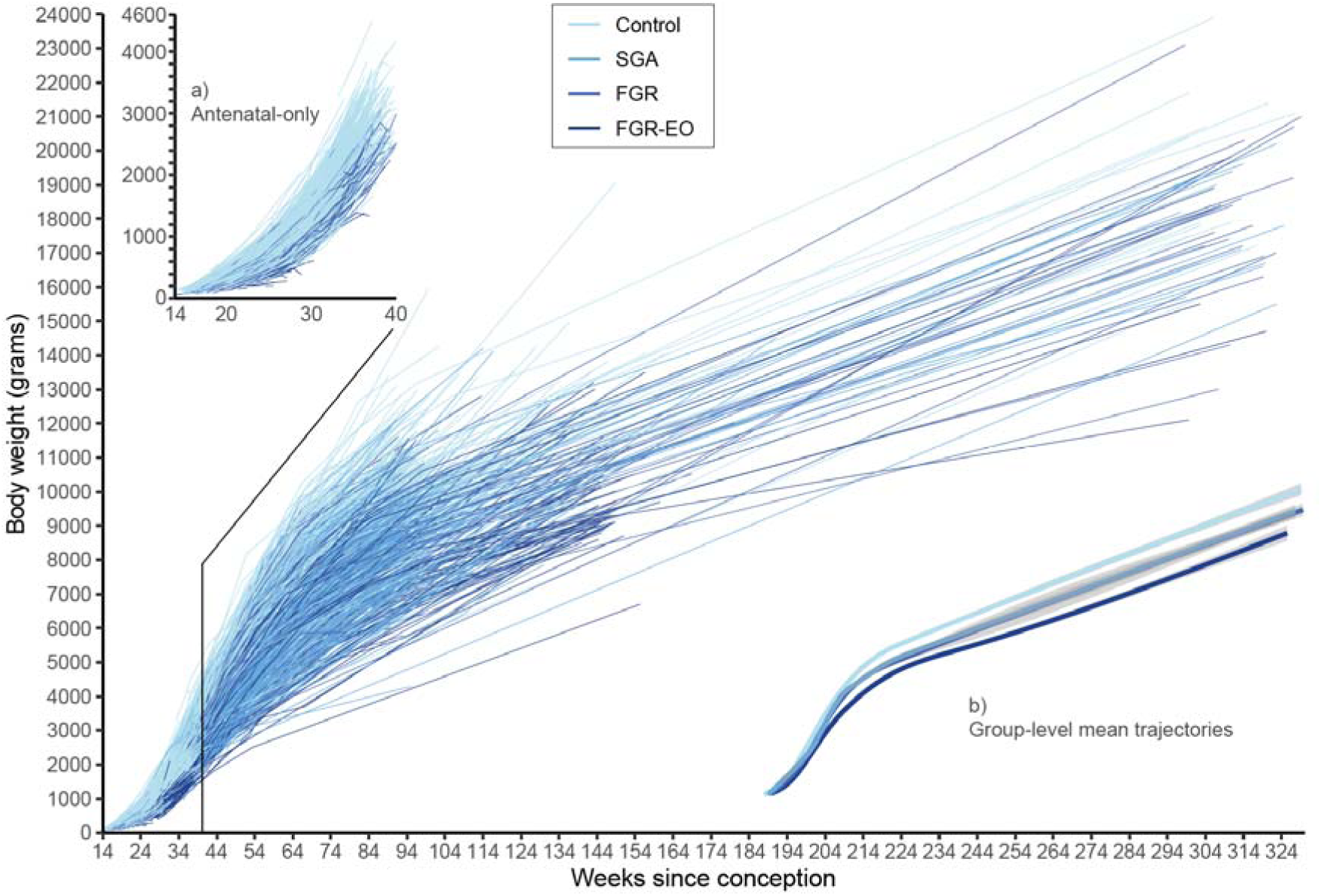
FGR and SGA subjects continued to have lower body weight to five years. Main: Subject-level body weight trajectories, pooled from the antenatal period (estimated fetal weight) through the postnatal period (actual weight), colour-coded by group. a) Plotting the antenatal measures alone confirmed the small fetal size expected in the non-control groups, most marked in FGR-EO. b) Group-level mean body weight trajectories, across the same developmental period as the main figure, allow to appreciate how all non-control groups continued to have lower mean body weight to five years; grey shading indicates standard error of the mean, note that the SGA and FGR mean trajectories are overlapping.

FGR and SGA subjects had lower weight at all intercepts examined, after adjusting for sex, antenatal vs postnatal at measure, weeks since conception at delivery, and data source (1666 subjects, 7402 measures (slightly smaller sample for statistical analysis, because not every antenatal data point was associated to weeks since conception at delivery)). For example, with the intercept anchored at 2 years, FGR-EO, FGR, and SGA subjects were 2176, 1270 and 1091 grams lighter than controls respectively (t = −28.688, t = −16.819, t = −18.004). In line with this, weight gain across the whole developmental period examined was slower in FGR-EO, FGR, and SGA subjects relative to controls by 15, 8, and 6 grams/week on average (t = −25.359, t = −12.095, t = −9.288).

### 3.6 FGR exacerbates the biological expense of growth

Previous work has quantified the biological expense of early-life growth by showing that higher body weight is associated with lower haematocrit (iron-containing red blood cells) levels ^31^. We investigated this in a subset of subjects for whom haematocrit level was measured at the same time as body weight (514 subjects, 730 measures; median 50 weeks since conception at measure (interquartile range: 43-92)). Median haematocrit level was 38%, in line with other published studies of this age range ^68^. Lower haematocrit level was predicted by higher body weight at the time of measure (−0.06%/100g, t = −2.983), and being FGR-EO or FGR (but not SGA) relative to controls (−4.70% and −2.26%, t = −2.677 and t = −2.381), after adjusting for sex, weeks since conception at delivery, and weeks since conception at measure (controls: 311 subjects, 453 measures; SGA: 156 subjects, 218 measures; FGR: 36 subjects, 45 measures; FGR-EO: 11 subjects, 13 measures).

### 3.7 Poor growth was associated with adverse motor and cognitive neurodevelopment

Finally, after demonstrating that FGR subjects struggled to gain weight at the group level, but to various degrees individually (Fig. 4 main), we tested whether this was associated with their neurodevelopmental scores. To do this, we entered each subject’s random effect from the growth model described in the final paragraph of section 3.5 as an explanatory variable. A more negative random effect indicates that the subject’s individual regression line was lower than predicted by the model, while a more positive random effect indicates that the subject’s individual regression line was higher than predicted by the model.

Motor score was 4 points lower in FGR subjects (FGR-EO t = −2.359, FGR t = −2.109) and 3 points lower in SGA subjects (t = −2.366), and also lower with more negative growth random effect (t = −3.730), after adjusting for sex, weeks since conception at delivery, and data source (1035 subjects). Cognitive score was 3 points lower in FGR-EO subjects (t = −2.136) (FGR and SGA not statistically significant), and also lower with more negative growth random effect (t = −2.534), after adjusting for the same covariates (1043 subjects). For motor score, there was no interaction between the effect of group and random effect, indicating that groups were similarly disadvantaged by poor growth. For cognitive score, growth had a stronger effect for FGR-EO (but not FGR or SGA) subjects than controls (interaction: t = 3.400). Given that the association between poor growth and adverse neurodevelopment affected all four groups, to visualise this we pooled groups and plotted mean body weight against weeks since conception, colour-coded by motor and cognitive outcome (Fig. 5).

**Fig. 5.**
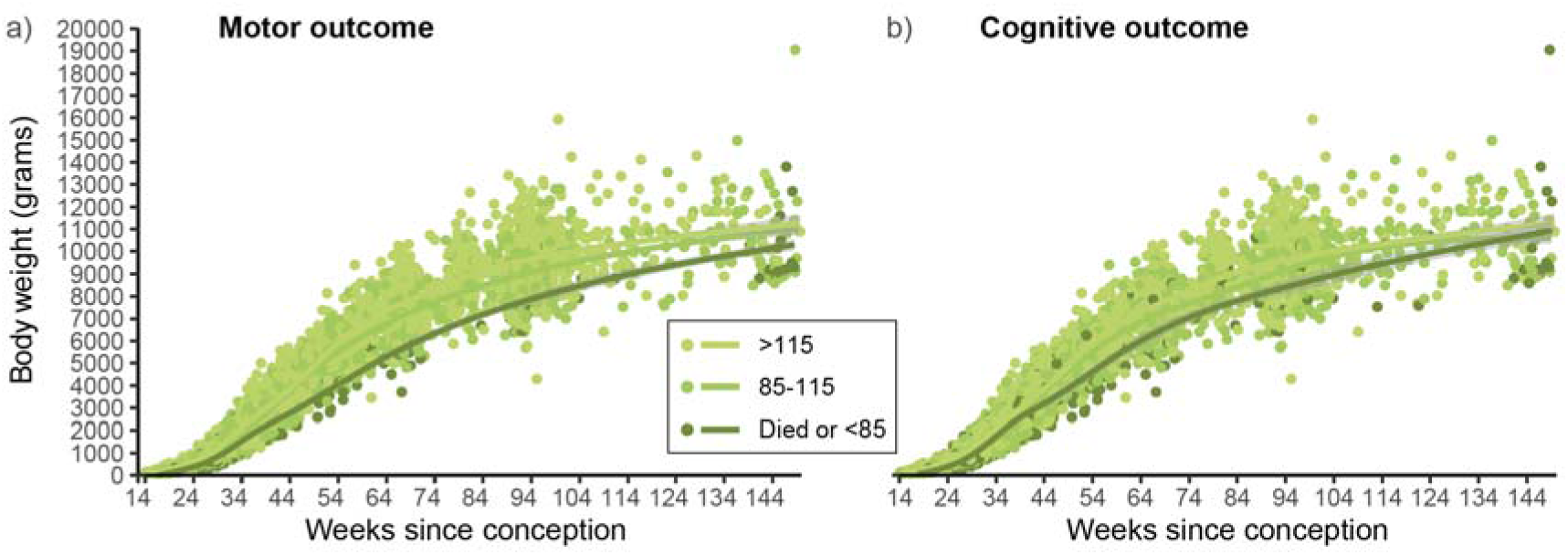
Lower body weight was associated with adverse motor (a) and cognitive (b) neurodevelopment. Mean body weight trajectories, colour-coded by outcome group. Note that the x axis stops at 2 years (150 weeks since conception) for consistency with the latest point that neurodevelopmental outcomes were assessed. Grey shading indicates standard error of the mean but is minimal when the data are dense and therefore difficult to appreciate; note in b) that the ‘85-115’ and ‘>115’ mean trajectories are almost totally overlapping so can barely be distinguished.

## 4. Discussion

In this study examining how metabolic differences develop, or manifest in growth trajectories, in fetuses and children from FGR, SGA, and normal pregnancies, our first finding is that FGR-EO subjects continue to exhibit higher whole-body metabolic rate for several weeks postnatally ^18,69^. Extrapolating from their perinatal heart rate being around 3.5 beats per minute faster, their heart will beat over 5000 times more per day across this period. This higher heart rate at rest may explain the origin of their dampened autonomic heart rate control, which is unmasked by a physiological challenge (clinically necessary heel lance) ^6,19^. Although previous studies have suggested that FGR infants do not respond to physiological challenges with the usual increase in heart rate, those stimuli lacked ecological validity (breathing CO, tilt table) ^69,70^. Here we show that FGR infants demonstrate weak heart rate reactivity to a naturalistic challenge that a hospitalised infant will regularly face. This complements our previous work showing that another group of vulnerable infants – those with higher physiological stress levels – also appear hypo-responsive to this same challenge ^71^. These data highlight the importance of accounting for inter-individual differences in physiological baseline, as infants maintain allostasis in a changing sensory environment.

As well as exhibiting evidence of higher whole-body metabolic rate (indexed by heart rate), our results imply that FGR subjects attempt to counteract this by minimising a major energy expenditure - brain metabolic rate - via reducing synapse number (indexed by white matter volume). Older FGR infants, at 1 year of age, also show reduced white matter volume, although with regional specificity^72^. In neonatal animal models, post-synaptic activity - the basis of the scalp EEG signal - is necessary for the development and maintenance of synapses ^73,74^, and we expected that attenuated EEG signal could be a mediator of lower synapse number in FGR. Although FGR infants did not, in fact, vary in the EEG metric assessed here, other work has noted differences in EEG activity in FGR subjects in line with this hypothesis, albeit with discrepancies across studies ^75^.

While FGR subjects adopt strategies to reduce energy expenditure, faltering growth indicates a residual shortfall. Here we demonstrate that fetal growth does not follow the usual law of association with placental size when that placenta is insufficient ^29^. Next, our results imply that this abnormal fetal growth reprogrammes growth into childhood, as infants are ill-resourced for postnatal weight gain and stay small until at least 5 years, failing to catch up with their peers even though they have left the compromised uterine environment. By stratifying the cohort into four groups, our growth curves visualise the increasingly negative impact on growth to five years of being SGA, FGR, or FGR-EO, relative to controls. In line with this, we highlight the strain of postnatal growth in FGR subjects especially, by showing how it depletes haematocrit from an already lower level. Finally, we model how poor growth in infancy is independently associated with adverse motor and cognitive outcomes ^51^, even after controlling for group and other risk factors like prematurity, with the effect most marked in FGR-EO subjects. In sum, we provide converging and multidimensional evidence of persistent and clinically significant metabolic differences after FGR.

This work has some limitations. The composite dataset was heterogenous, but allowed the largest study - to our knowledge - which includes longitudinal fetal-postnatal courses associated to FGR. However, while the majority of analyses benefited from large sample sizes, other data samples were modest, including for the heart rate reactivity and EEG parts. In addition, socio-economic information was not included here, but is likely to have contributed to neurodevelopmental outcomes ^43^. Further, doppler ultrasound information was not quantitatively examined, because this was unavailable for many data points, but this is the gold standard method to assess FGR and its severity ^48^. Finally, future work should account for fetal conditions which commonly present alongside FGR but could affect neurodevelopment in their own right, like oligohydramnios (reduced amniotic fluid) which will affect the fetal sensory environment.

In conclusion, this comprehensive study of different developmental domains (cardiac, autonomic, neurological, physical, haematological, behavioural) shows the complex effects of FGR, which are uniquely evident when integrated together within a unified metabolic framework. Together, our results represent FGR as an evolving neurological compromise which reaches far into the postnatal period, and in which metabolic and autonomic adaptations to the initial insult may further harm development, beyond the initial antenatal insult. Growth trajectories encode information about how FGR is transmitted into adverse neurodevelopmental outcomes, and could identify intervention opportunities in this vulnerable population.

## Data Availability

The EEG data described will be made accessible on Open Science Framework at publication. For the three open-access datasets, we will provide the reference numbers of the participants analysed in a supplementary spreadsheet, for replication purposes. Please direct enquiries about any other data to the corresponding author.

## Acknowledgements

We warmly thank the women and families who contributed to this research. KW acknowledges helpful discussions about these data with Professor Van Savage, Aisling Roya-Murran, and Professor Samraat Pawar, and support from Dr Judith Meek regarding clinical aspects of data collection at University College London Hospitals (UCLH). We also acknowledge Prof A David Edwards and the rest of the ePrime team with thanks. This work was primarily supported by Brain Research UK (awardee: KW). BC’s internship with KW, during which BC worked on these data, was supported by a Wellcome Biomedical Vacation Scholarship. LF is supported by the Medical Research Council (MR/X010716/1). We acknowledge the support of the National Institute for Health Research UCLH Biomedical Research Centre. The EVERREST study received funding from the European Union Seventh Framework Programme (FP7/2007-2013) under grant agreement no. 305823, the Rosetrees Trust and the Mitchell Charitable Trust in memory of Shoshana Mitchell Glynn. We acknowledge the contribution of the EVERREST consortium members:

- UCL: Rebecca Spencer, Kasia Maksym, Yuval Ginsberg, Tal Weissbach, Donald M Peebles, Ian Zachary, Neil Marlow, Gareth Ambler, Anna Morka, Jade Dyer, Helen Knowles, Steve Hibbert, Kate Maclagan, Carlo Rossi, Neil J Sebire
- UCLH: Angela Huertas-Ceballos, Gina Buquis, Jade Okell, Ingran Lingam
- Department of Obstetrics and Fetal Medicine, University Medical Center Hamburg-Eppendorf, Hamburg, Germany: Kurt Hecher, Anke Diemert
- Department of Obstetrics and Gynaecology, Institute of of Clinical Sciences Lund, Skane University Hospital, Lund University, Lund, Sweden: Karel Maršál, Stefan R. Hansson, Jana Brodszki
- Institut D’Investigacions Biomèdiques August Pi í Sunyer, University of Barcelona, Barcelona Center for Maternal-Fetal and Neonatal Medicine, Barcelona, Spain: Francesc Figueras, Eduard Gratacós

