## Supplementary material for "Metabolic expenditure, neurodevelopment, and weight gain into early childhood after fetal growth restriction": Table S1

**Supplementary Methods**

**Table S1: Details about each data source**

|  | HR | HR reactivity | MRI | EEG | Body weight | Placenta  weight | Haem | Bayley scores | Site | Subs |
| --- | --- | --- | --- | --- | --- | --- | --- | --- | --- | --- |
| UCLH | Yes | Yes | No | Yes | Yes | Yes | No | Yes | NNU | FGR-EO, FGR, SGA, Controls |
| EVERREST | Yes | No | No | No | Yes | Yes | No | Yes | AN | FGR-EO |
| e-Prime | No | No | Yes | No | Yes | No | No | Yes | NNU | FGR-EO, Controls |
| FEMINA | Yes | No | No | No | Yes | No | No | No | AN | FGR-EO, FGR, SGA, Controls |
| IEEE | Yes | No | No | No | No | No | No | No | AN | FGR, Controls |
| Norway | No | No | No | No | Yes | No | No | Yes | AN & NNU | FGR-EO, FGR, SGA, Controls |
| Alabama | No | No | No | No | Yes | Yes | Yes | Yes | AN & NNU | FGR-EO, FGR, SGA, Controls |

HR = Heart rate. Haem = haematocrit. Site = Site of recruitment. NNU = Neonatal unit. AN = Antenatal. Subs = Subgroups
